# Optimal vaccination in aging populations under age-dependent infection fatality risks

**DOI:** 10.64898/2026.06.24.26356423

**Authors:** Michiel van Boven, Christiaan H. van Dorp, Maikel Bosschaert, Jurjen van der Schans, Debbie van Baarle, Mirjam E. E. Kretzschmar

## Abstract

**Background:** Vaccination programs have greatly reduced the burden of infectious diseases, particularly in childhood. As populations age, however, the burden of respiratory infections such as influenza A, respiratory syncytial virus (RSV), and SARS-CoV-2 increasingly falls on older adults. Because infection fatality rates rise steeply with age, vaccination strategies that alter the age distribution of infections may have complex population-level consequences. We used transmission models to examine how the timing and frequency of vaccination influence infection-induced mortality and years of life lost (YLL) in aging populations.

**Methods and findings:** We analyzed age-structured transmission models that incorporate demographic change, age-specific infection fatality rates, and waning immunity after infection or vaccination. We varied the age at first vaccination, vaccination intervals, and coverage across a wide range of pathogen characteristics, including transmissibility and the duration of natural and vaccine-induced immunity. For single-dose vaccination programs with long-lived protection (5–50 years), the age at vaccination minimizing mortality in older adults for pathogens with strongly age-increasing fatality risk typically ranges from 60 to 80 years. The optimal age shifted toward older ages when transmissibility was higher or natural immunity lasted longer. Repeated vaccination produced qualitatively different outcomes. When vaccine-induced immunity was short-lived (*<*5 years), vaccination can shift infections toward the oldest ages where fatality risks are highest, increasing both mortality and YLL compared with no vaccination. This study has limitations. Our analysis used stylized transmission models and assumed vaccines that fully prevent infection, which may overestimate age-shifting effects compared with real-world vaccines that primarily reduce disease severity.

**Conclusions:** Optimal adult vaccination strategies depend jointly on pathogen transmissibility, the duration of immunity, and population demography. Vaccination programs that suppress infections earlier in life without protecting individuals into late life may shift infections toward ages with higher fatality risk. These findings highlight the need to evaluate adult vaccination strategies across the full life course and have important implications for vaccination policies against influenza A and other pathogens with strongly age-dependent infection fatality rates.

**Author Summary:** *Why was this study done?:* Populations worldwide are aging rapidly, increasing the burden of respiratory infections such as influenza A, respiratory syncytial virus (RSV), and SARS-CoV-2 in older adults. Vaccination of adults is therefore becoming an increasingly important public health strategy. However, there are no general principles for deciding at which ages vaccination should be offered.

*What did the researchers do and find?:* We used mathematical transmission models to examine how the timing and frequency of vaccination influence infection-related mortality in aging populations. We found that the optimal age for vaccination depends strongly on pathogen characteristics such as transmissibility and the duration of immunity. In some situations, vaccination can even increase mortality by shifting infections to older ages where fatality risks are highest.

*What do these findings mean?:* These results show that there is no universal rule for the optimal timing of adult vaccination. Vaccination strategies should account for how vaccination changes the age distribution of infections over the life course. Our findings provide guidance for designing vaccination strategies against influenza A and other pathogens in aging societies.

## Introduction

Populations worldwide are aging rapidly due to sustained declines in mortality and fertility [46, 40]. This has profound consequences for public health, as older individuals are generally more frail and more likely to suffer from severe outcomes of infections that are usually mild in younger age groups. For influenza A, infection fatality rates are very low below 60 years but rise steeply thereafter, in both low- and high-risk persons [8, 2]. Likewise, for SARS-CoV-2, infection fatality rates increase approximately log-linearly with age [27]. These patterns imply that the demographic transition to populations dominated by older adults can substantially increase the burden of infection-induced mortality over the coming decades.

Childhood immunization programs have strongly reduced the infectious disease health burden since their introduction in the 1950s. In contrast, vaccination of older adults has gained attention relatively recently, driven by demographic shifts and the rising impact of respiratory pathogens in older populations [23, 22, 42, 10, 2]. Yet, current assessments of adult vaccination policies rarely account for the combined effects of demographic change, contact patterns, and waning immunity. In particular, little is known about how the interplay between short-lived vaccine protection and longer-lasting natural immunity may shift infections toward older ages. This may have unintended effects on infection-induced mortality if the infection fatality rate increases with age.

Here, we investigate the optimal timing of vaccination using age-structured transmission models that incorporate demographic projections, age-specific infection fatality rates, empirical contact patterns, and waning immunity. Motivated by respiratory pathogens such as influenza A, respiratory syncytial virus, and SARS-CoV-2, we vary key parameters such as pathogen transmissibility (*R*_0_), vaccination coverage, age at first vaccination, and intervals between vaccinations. A central assumption is that the infection fatality rate increases steeply with age [20, 3]. Taking a long-term demographic perspective, we determine the number of infection-induced deaths and years of life lost (YLLs) to identify vaccination schedules that minimize the long-term burden of disease.

We show that outcomes of adult vaccination can be highly non-trivial. Specifically, depending on the balance between aging, transmissibility, and waning immunity, vaccination can either markedly reduce or inadvertently increase infection-induced mortality. For repeated yearly vaccinations, mimicking influenza A, such an adverse outcome is most likely when transmissibility is high and vaccine protection is short-lived relative to infection-induced immunity, thereby shifting infections to older ages. Optimal vaccination schedules therefore cannot be inferred from pathogen severity or transmissibility alone but depend on demographic and immunological dynamics.

## Results

### Demography

Population aging is incorporated using age- and time-specific mortality projections for the Netherlands from the Royal Dutch Actuarial Association [19, 26]. These projections are based on the Li-Lee mortality model and are calibrated to mortality data from 19 European countries [21]. Here, we approximate the projected mortality rates for ages 15 years and older with a log-linear function that has been fitted to the projections for the Netherlands,

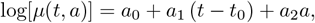

where *t*_0_ = 2020 (*t* ≥ *t*_0_), and *a*_0_ = −11.64, *a*_1_ = −0.014, and *a*_2_ = 0.11 are the estimated parameters. These estimates yield mortality rates that are close to projections for other high-income western European countries [16].

Figure 1 shows the survival projections with the fitted log-linear model (left-hand panel) and shows the corresponding survival probabilities by age and year (right-hand panel). By not taking into account time-age interactions, the fitted function slightly underestimates mortality below age 30 years and overestimates it beyond age 105 years. The resulting mortality patterns closely match those observed in other high-income countries, particularly in Western Europe [19], and the log-linear specification can be readily adapted to alternative demographic scenarios. Throughout, remaining life expectancy and years of life lost (YLL) by infection are calculated using cohort life expectancy at a given age, thus taking into account that age-specific mortality rates tend to go down over time as persons age. Notice that at any age cohort life expectancy is expected to increase over time such that, for instance, remaining life expectancy at 75 years was 12.7 years in 2020, 16.8 years in 2060, and 21.3 years in 2100. These values are based on the estimated log-linear mortality rate, and are close to estimates based on the actual projected mortality rates.

**Figure 1.**
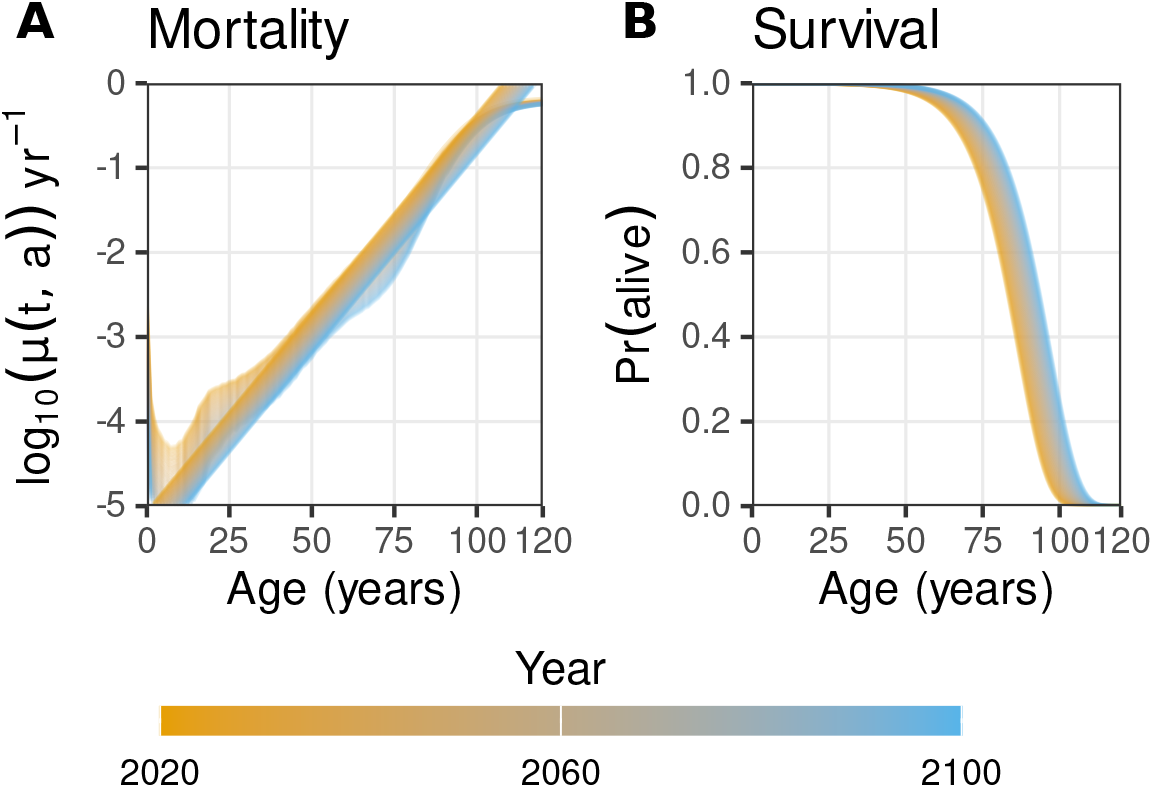
Projected mortality rates and survival probabilities by age and time. The left-hand panel shows the projected age- and time-specific mortality for the Netherlands (curved lines) together with the fitted mortality (straight lines). The right-hand panel shows the corresponding survival probabilities as a function of age.

Besides mortality, the demography of the host population is shaped by recruitment through births and net migration. To retain generality and to prevent making the analyses needlessly Netherlands-centric, we use a stylized demographic set-up in which the Netherlands serves as a baseline. In all simulations, the initial population composition is constructed using the projected Dutch mortality rates for 2025. Because future birth and migration rates are difficult to forecast and vary widely across countries, we assume a fixed annual birth cohort of 200,000 individuals and set migration rates to zero.

### Single vaccination

To set the stage, we consider a stylized scenario with a single vaccination per lifetime, at age *a*_v_. Demography is kept fixed as described above (baseline age structure calibrated to projected Dutch mortality in 2025, with constant recruitment through births), such that differences between scenarios are driven by transmission and immunity rather than by specific demographic details.

Figure 2 summarizes the resulting mortality impact relative to the corresponding no-vaccination baseline. Across immunity assumptions, one-off vaccination yields the largest reductions in mortality when transmission is weak (low ℛ_0_). When immunity is short-lived (panels A–B), vaccination is most effective when administered at older ages, and the vaccination age that minimizes mortality increases with ℛ_0_. In contrast, when immunity is longer-lasting (panel C), the optimum shifts towards vaccinating young children, consistent with maximizing indirect protection via herd effects. However, for sufficiently large ℛ_0_ combined with long-lasting immunity (panel D), one-off vaccination can become counterproductive, with mortality increasing relative to no vaccination, reflecting that partial suppression of early infections may primarily postpone infections to older ages where outcomes are worse.

**Figure 2.**
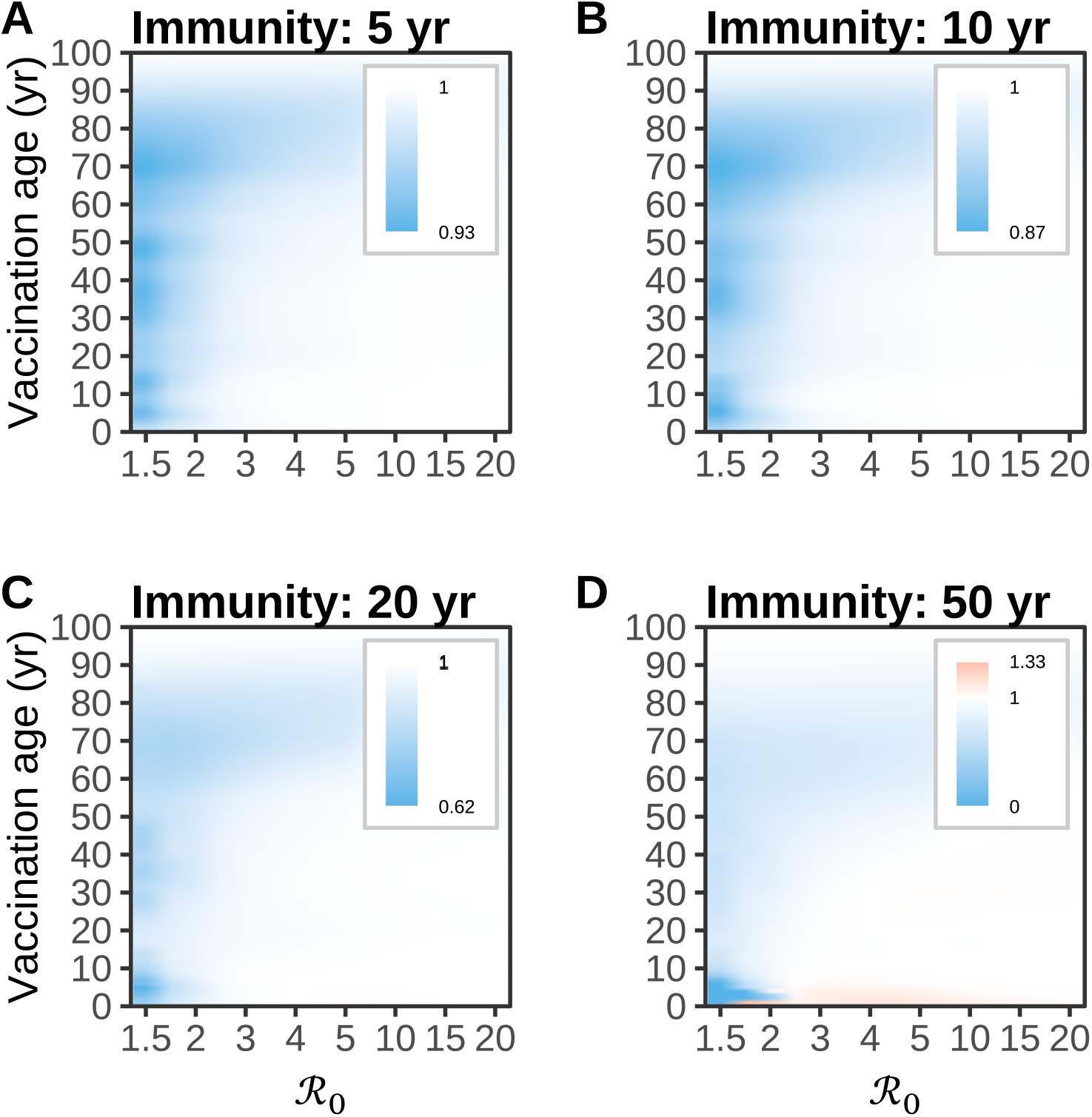
Relative mortality as a function of pathogen transmissibility (ℛ_0_) and age at vaccination. For given ℛ_0_ relative mortality is calculated as the ratio of the infection fatality rates in scenarios with and without vaccination. The duration of vaccine induced immunity is set at half the indicated duration of natural immunity (i.e 2.5, 5, 10 and 25 years). A single vaccination dose is given at the indicated vaccination age and vaccination coverage is set at 80%.

The heatmaps in Figure 3 make this age pattern explicit by showing the optimal vaccination age as a function of ℛ_0_ and the immune period, both when minimizing mortality and when minimizing years of life lost (YLL), and under two mortality schedules (2025 versus 2050). For short immune periods, the optimum lies consistently at older ages and increases with ℛ_0_ (typically ~70–85 years for mortality minimization). For long immune periods and low ℛ_0_, the optimum lies in early childhood (near 0–5 years), whereas for higher ℛ_0_ the optimum shifts abruptly to late-life vaccination (around 60–70 years for mortality and somewhat younger for YLL), mirroring the regime where childhood vaccination becomes less beneficial or harmful due to age-shifting of infections. These qualitative patterns are robust to whether the baseline mortality schedule corresponds to 2025 or 2050.

**Figure 3.**
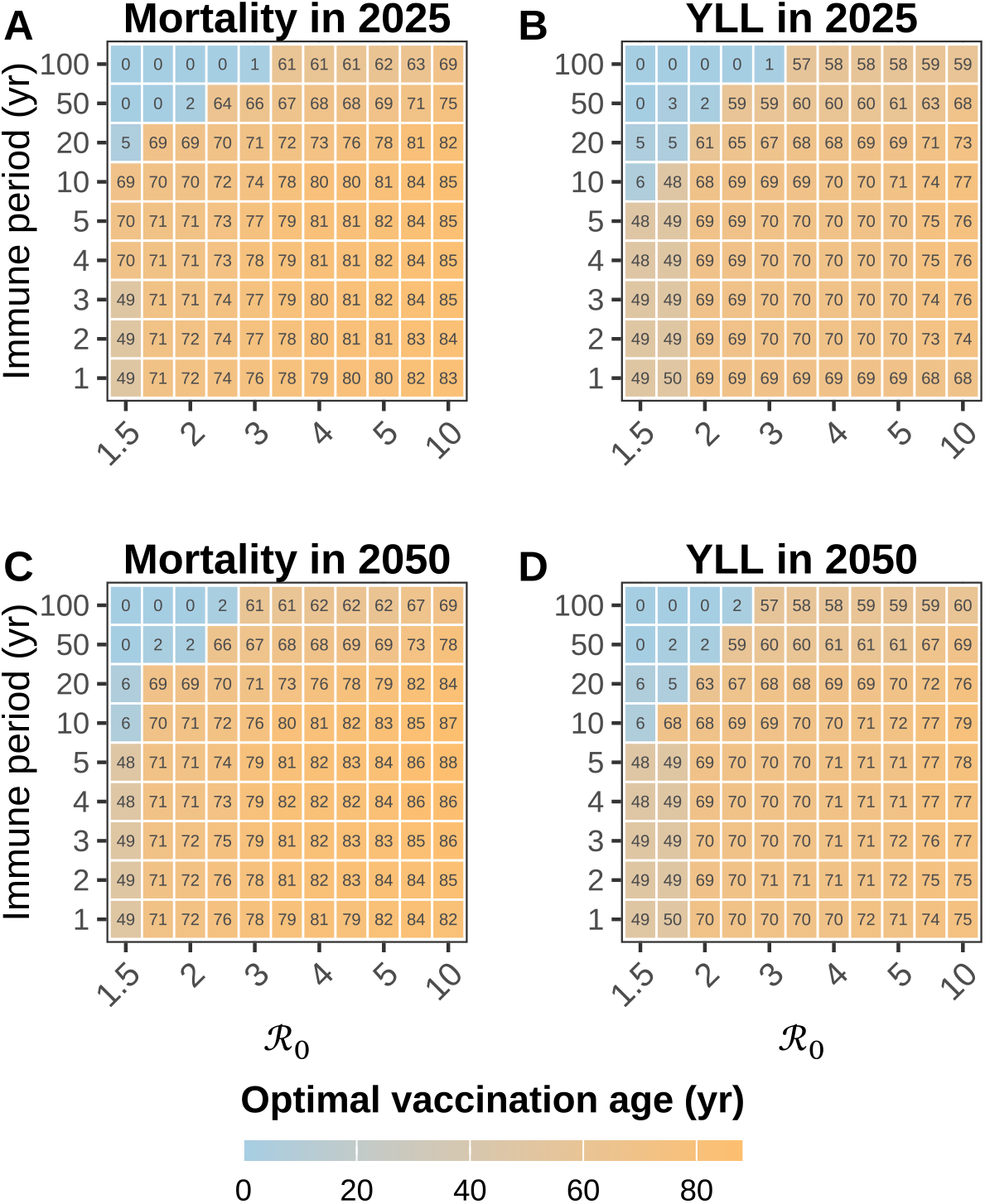
Optimal age at vaccination as a function of the basic reproduction number (ℛ_0_) and the duration of natural immunity. **(A)** Age minimizing total mortality in 2025. (**B**) Age minimizing total years of life lost (YLL) in 2025. (**C**) Age minimizing total mortality in 2050. (**D**) Age minimizing total years of life lost (YLL) in 2050. Each cell represents results from 101 independent simulations, corresponding to vaccination started at ages 0 through 100 years. Every scenario assumes a single vaccination event per individual, without revaccination, and a fixed demographic structure corresponding to the year 2025. The duration of immunity on the horizontal axis refers to naturally acquired immunity, and vaccine-induced immunity is assumed to last half as long.

### Repeated vaccination

Motivated by influenza A we next consider repeat vaccinations, assuming that both natural and vaccine-induced protection against infection are relatively short-lived. Vaccination is administered yearly from a certain age onward in an October–November window, mimicking actual influenza vaccination campaigns. By default we assume an effective vaccination coverage of 50% [12, 28]. To further increase realism we assume weak seasonal forcing (5%) [36], while outcomes are summarized over a long horizon of 50 years to reduce the impact of differences between influenza seasons caused by stochastic variation in waning rates (see Materials and Methods for details). Specifically, naturally acquired immunity is allowed to wane on a multi-year time scale (varied between 2–7 years across scenarios), whereas vaccine-induced immunity is short-lived (ranging from 0.5-5 years) [13, 45, 43]. Unless stated otherwise, demography is fixed as projected for 2025, but with realistic cohort life expectancy as in Figure 1, so that differences between scenarios cannot be caused by changing population structure.

Figure 4 shows a single simulation run with ℛ_0_ = 3, effective vaccination coverage of 80%, duration of vaccine protection of 2 years, and vaccination offered from the age of 60 onward. Panel A shows how repeated campaigns generate a large persistent vaccinated compartment, and dampen the number of infections. Panels B and C show the corresponding age–time heatmap of the prevalence of infection and age-aggregated mortality. Overall, we find that, even when seasonal forcing is weak, there are pronounced and highly variable epidemics. These epidemics give rise to highly variable yearly mortality.

**Figure 4.**
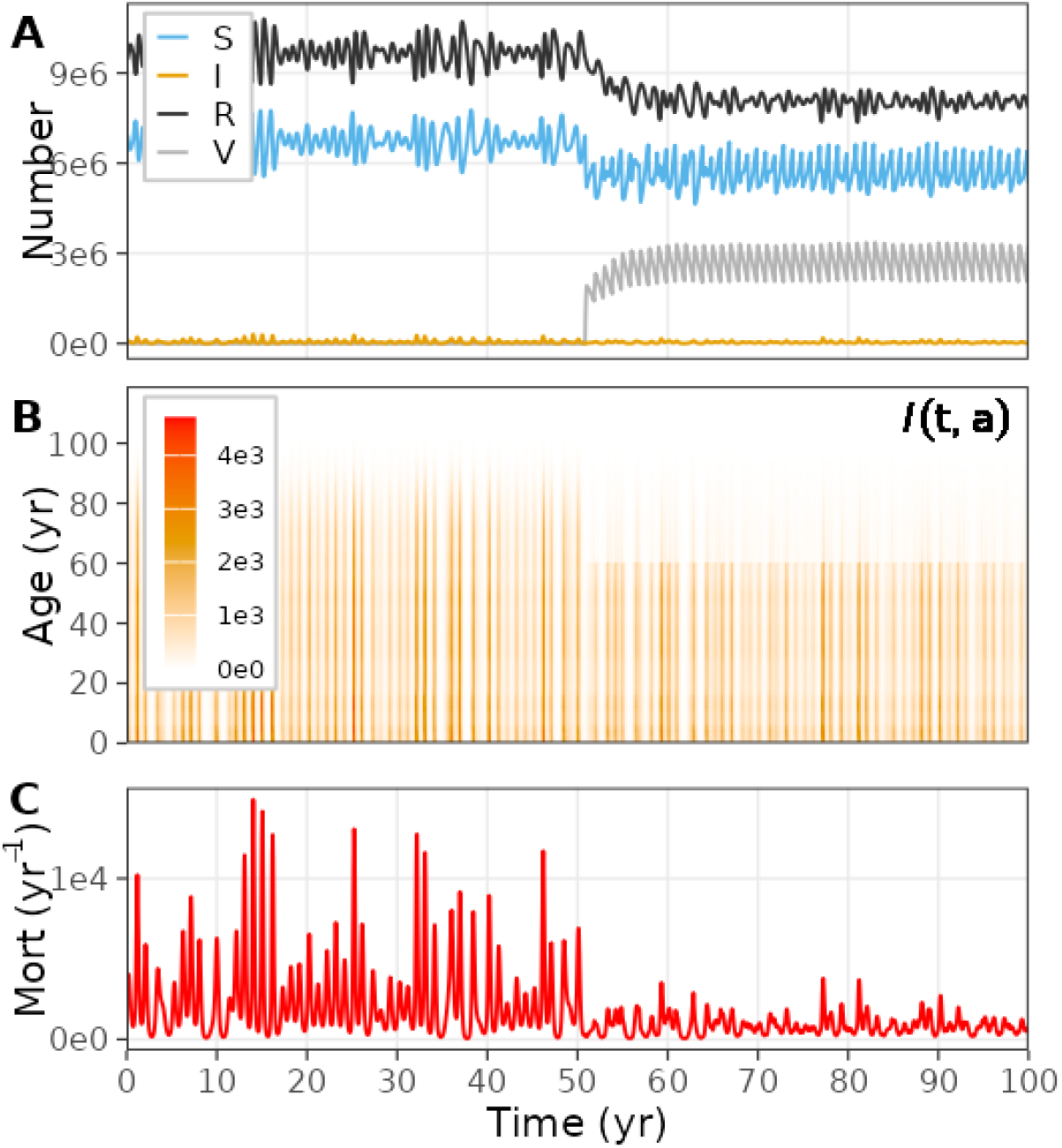
Example dynamics under repeated annual vaccination. Shown is a representative simulation with vaccination started at *t* = 50, and with ℛ_0_ = 3, vaccination coverage of 80%, duration of vaccine protection of 2 years, weak seasonal forcing (5%), and yearly vaccination administered from the age of 60. (A) Time series of compartment sizes (susceptible *S*, infected *I*, recovered *R*, vaccinated *V*). (B) Age–time heatmap of the infection prevalence. (C) Mortality rate over time, illustrating highly variable episodic mortality peaks despite weak forcing.

To investigate the impact of repeated vaccination systematically, Figure 5 shows mortality relative to no vaccination as a function of pathogen transmissibility (as measured by the basic reproduction number ℛ_0_) and the age at first vaccination, for different durations of vaccine-induced immunity (0.5, 1, 2 and 5 years). To do so in a semi-realistic manner, we connect ℛ_0_ to empirically reported early-season effective reproduction numbers (*R*_*e*_) by writing *R*_*e*_ = ℛ_0_ *S*_eff_ (0), where *S*_eff_ (0) is an effective susceptibility summarizing the reduction in transmission potential at the start of the season due to pre-season immunity and other unmodeled factors (e.g., heterogeneity in risk and mixing). Estimates based on early epidemic growth commonly place influenza A effective reproduction numbers around *R*_*e*_ ≈ 1.1–1.6 across countries and seasons [37]. Joint inference studies that estimate both susceptibility and transmissibility further suggest *S*_eff_ (0) ≈ 0.65–0.71 with ℛ_0_ ≈ 1.86–2.04 for US influenza seasons, implying early-season *R*_*e*_ in the range ≈ 1.18–1.40 [44]. Because our model uses a constant ℛ_0_ that does not include seasonal forcing, and as there is substantial uncertainty about which mechanisms dominate observed seasonal patterns [38], we vary ℛ_0_ broadly (1.5–5) to span realistic early-season *R*_*e*_ values under plausible *S*_eff_ (0). For example, *R*_*e*_ = 1.1–1.6 corresponds to ℛ_0_ = *R/S*_eff_ (0) in the range 1.4–5.3 when *S*_eff_ (0) ∈ [0.3 − 0.8].

**Figure 5.**
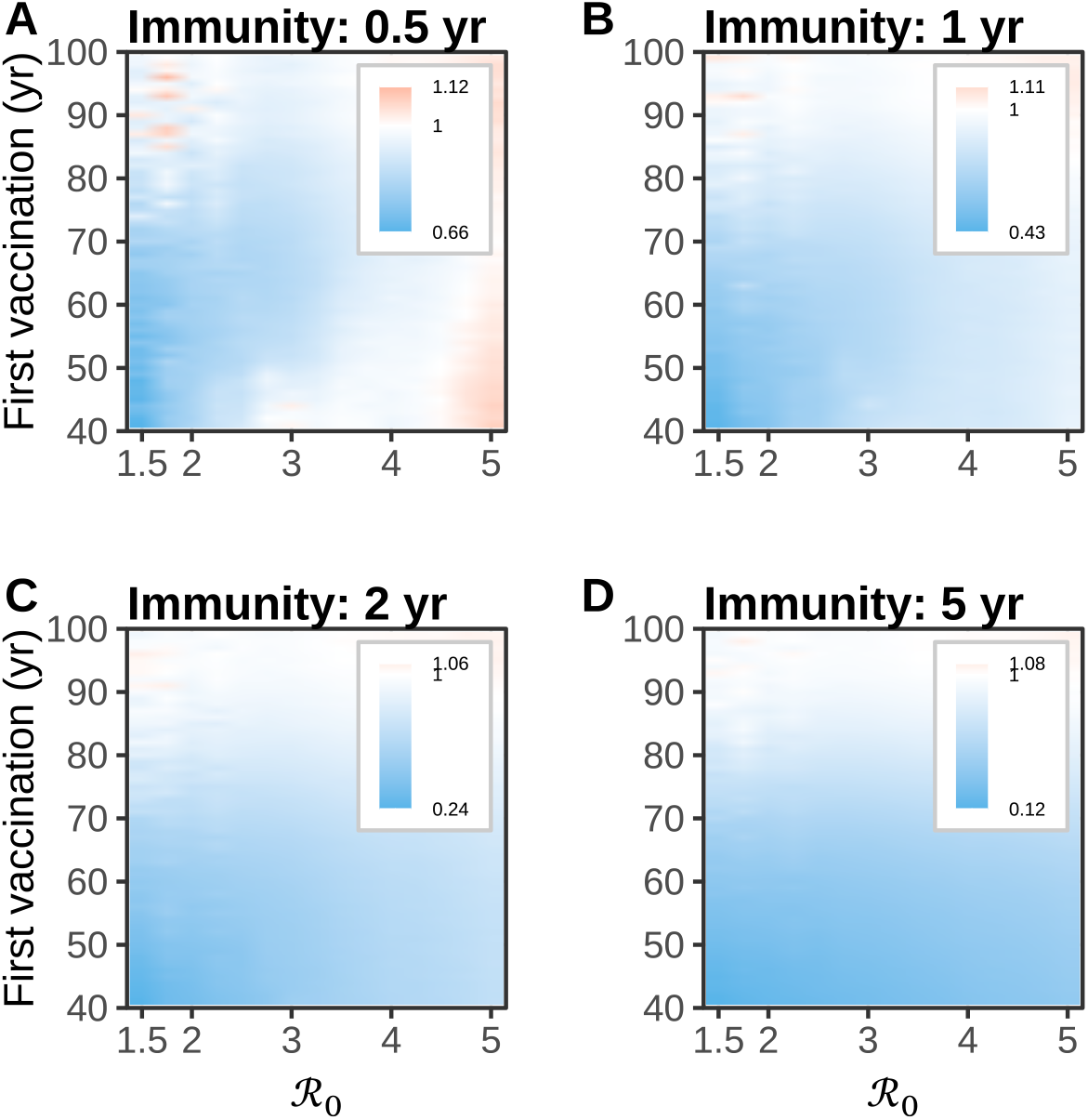
Impact of repeated annual vaccination on mortality in an influenza-like scenario. The heatmaps show mortality under vaccination relative to no vaccination as a function of pathogen transmissibility (ℛ_0_) and the age at which vaccination is first offered. Panels correspond to different durations of vaccine-induced protection (0.5, 1, 2, and 5 years). In all scenarios vaccination is applied annually within a fixed seasonal window (October–November), with campaign coverage of 80%, weak seasonal forcing (5%), vaccination continued up to age 100 years, and naturally acquired immunity varying between seasons from 2–7 years). Values below 1 indicate reductions in mortality, whereas values above 1 indicate increased mortality relative to no vaccination.

When transmissibility (ℛ_0_) is low, starting vaccination at younger ages yields the largest mortality reductions, consistent with indirect protection (i.e. herd immunity effects) accumulating over repeated rounds. For higher ℛ_0_, starting vaccination early remains beneficial only when vaccine-derived immunity is sufficiently long-lived (notably the 2–5 year panels), where repeated vaccination can both protect vaccinated persons and reduce transmission enough to lower mortality in older ages.

A qualitatively different regime emerges when transmissibility is high and vaccine immunity is short-lived (0.5–1 year) [13, 45]. In this setting, starting vaccination at young ages can increase mortality above the no-vaccination baseline (red regions in Figure 5), because vaccination reduces or delays infections at younger ages without providing sustained protection into late life. As a consequence, infections are shifted to older ages where the infection-fatality risk is much higher. This perverse outcome is amplified by the steep age gradient in the infection fatality rate, rising exponentially with age [34, 41, 33, 20]. At the same time, remaining life expectancy falls rapidly with age, so that postponing infections from midlife to older ages both increases the per-infection fatality risk and reduces the scope for subsequent life years to be saved (i.e. a competing-risk effect). Finally, starting vaccination only at older ages is generally ineffective in this repeated-vaccination setting as any reduction in transmission is too small and too late to offset the tendency of vaccination to shift infections upward in age.

Finally, we analyze a scenario with lower effective vaccination coverage of 30%. This models the situation with vaccination coverage of 50% and vaccine effectiveness of 60%, so that in effect 0.6 *×* 50% = 30% of the population is protected [29, 4, 31, 17, 5]. Figure 6 repeats the analysis of Figure 5 with the lower effective vaccination coverage. The overall structure of the heatmaps is preserved, but mortality reductions are attenuated across the parameter space, consistent with weaker direct protection and weaker indirect benefits. Conversely, the region in which repeated vaccination increases mortality expands, most notably when vaccine-induced immunity is short-lived (0.5–1 year) and transmissibility is high, because partial suppression of infections is more likely to shift residual risk towards older ages.

**Figure 6.**
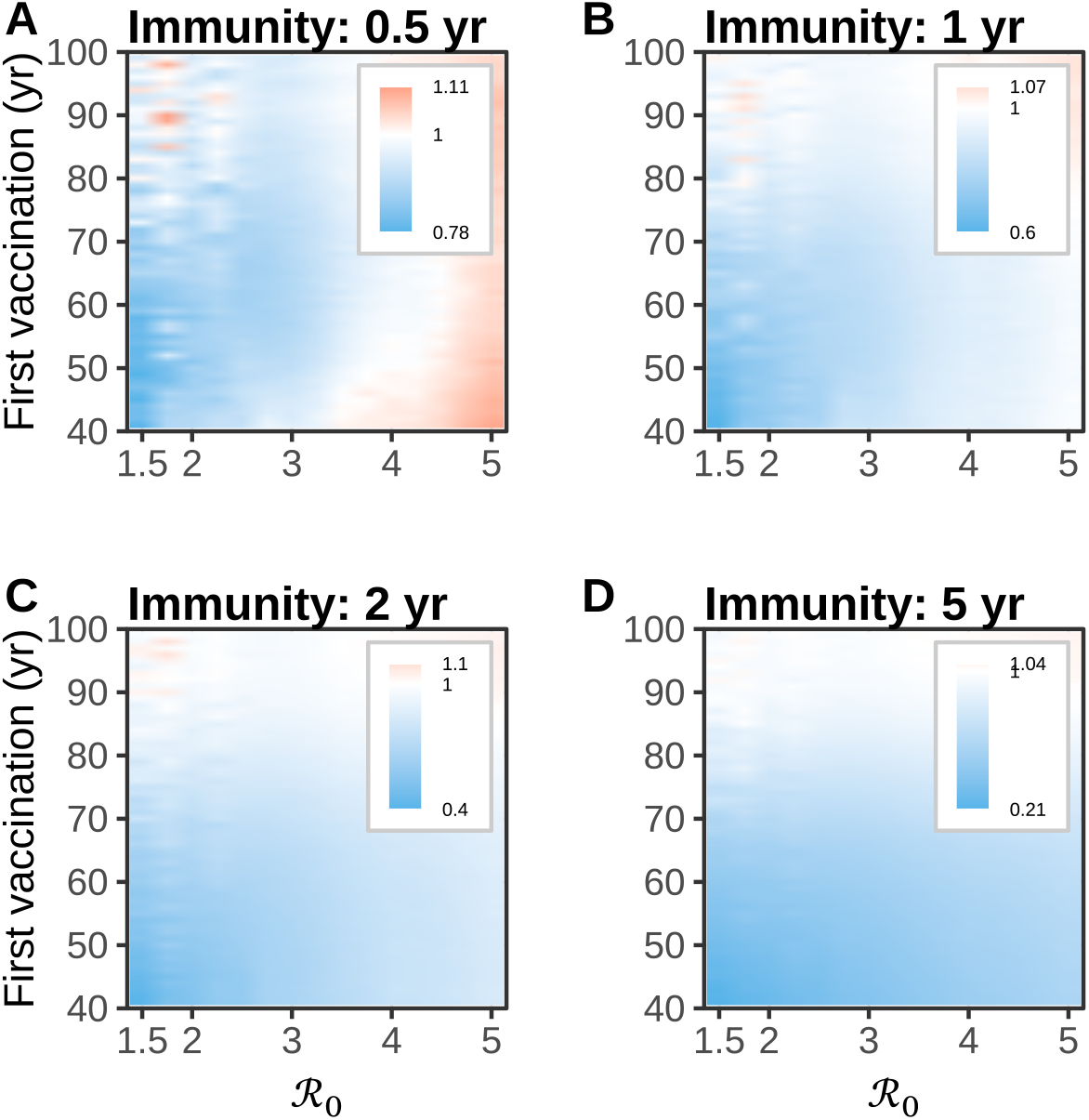
Impact of repeated annual vaccination on mortality in an influenza-like scenario with low effective vaccination coverage (30%). The heatmaps show mortality under vaccination relative to no vaccination as a function of pathogen transmissibility (ℛ_0_) and the age at which vaccination is first offered. Panels correspond to different durations of vaccine-induced protection (0.5, 1, 2, and 5 years). For details, see Figure 5. Values below 1 indicate reductions in mortality, whereas values above 1 indicate increases relative to no vaccination.

## Discussion

We showed that in aging populations, the optimal timing of vaccination depends critically on pathogen transmissibility and the duration of immunity and cannot be inferred from pathogen severity alone. Specifically, we analyzed how vaccination should be timed in aging populations when the infection fatality risk increases steeply with age, using influenza A as our main example. Two general messages have emerged. First, even in a relatively simple age-structured transmission model, there is no single best age to vaccinate older adults, as the optimal timing depends jointly on pathogen transmissibility, the duration of naturally acquired immunity, and the duration of vaccine-derived immunity. Second, repeated vaccination can have qualitatively different and sometimes even counterintuitive population-level effects. Specifically, when vaccination suppresses infection at relatively young age (under 60 years, say) without providing sustained protection into late life, it tends to postpone infections to ages with the highest mortality rates. This could offset some or even all of the direct gains.

Our single-vaccination analyses provide a baseline for understanding why optimal ages cluster in late adulthood under many plausible scenarios. When immunity is short-lived and the pathogen is highly transmissible, the indirect benefits of vaccinating early are limited. In that regime, the dominant factor is direct protection at ages where the hazard of a severe outcome rises sharply. As a result, the mortality-minimizing age tends to lie in the 60–80 year range and increases with ℛ_0_ (Figure 3, Figure 5). In contrast, when immunity is long-lived and transmissibility is low to intermediate, vaccinating at younger ages can generate substantial indirect protection via herd immunity. The sharp regime shifts we observe reflect a tension between (i) vaccinating early to suppress transmission and (ii) vaccinating late to protect when the consequences of infection become most severe, while taking into account population demography.

A natural question is how our conclusions depend on infection-induced mortality and demographic composition. In our model, these factors turned out to have only a modest influence on the optimal vaccination age. As shown in Figure 3, changing background mortality and population age structure from 2025 to 2050 shifts the optimum vaccination age upward by only 1–3 years. Furthermore, our results are driven primarily by relative differences in risk across ages and by the steepness of the age gradient of the infection fatality rate, rather than by the absolute scale of the mortality function. In fact, multiplying ifr(*a*) by a constant factor scales deaths and years-of-life lost (YLL) almost proportionally, but does not materially change which vaccination age is optimal. By contrast, the choice of outcome measure matters more. Mortality minimizing optima are often around 80 years, whereas YLL-minimizing optima are often around 70 years. This is caused by the fact that YLL places more weight on preventing deaths at younger ages, where remaining life expectancy is higher.

Repeated vaccination adds realism to our model. Still, the qualitative patterns remain similar, and we observed the same trade-off between direct protection and shifts in the age distribution of infections. In practice, however, the consequences of repeated vaccination are almost invariably assessed on a per-season basis. As we have shown, this can be problematic. First, starting vaccination at old age can be inefficient, because remaining life expectancy is short. Second, and more importantly, expanding eligibility to younger ages can increase long-run mortality when transmissibility is high and the duration of vaccine-derived protection is short relative to infection induced protection (Figure 5). This can be explained as follows. Vaccination can suppress or delay infections in midlife, but if protection does not extend reliably into late life, a larger share of infections can occur at ages with much higher infection fatality risk. This is a population-level version of the well-known phenomenon that partial suppression of infection can increase the average age at infection, with potentially negative consequences when severity rises with age. A well-known real-world example is the increase in congenital rubella syndrome in the early 1990s following the introduction of rubella vaccination in Greece in the mid-1970s [30].

Considering the above, how realistic is a perverse outcome for influenza A or other respiratory pathogens? Several features of influenza A epidemiology and vaccination practice could dampen the mortality-increasing regimes of our models. First, influenza vaccines are often imperfect at preventing infection, while still reducing severe outcomes [4, 14, 7, 32]. Hence, a vaccine that primarily reduces severity rather than susceptibility produces less age-shifting of infections than the all-or-nothing vaccine assumption used here. Second, naturally acquired protection against influenza infection is complicated by antigenic drift and immune history, which can shorten the effective duration of protection against currently circulating strains and thereby reduce the contrast between vaccine- and infection-induced immunity. Third, actual vaccination programs do not achieve uniformly high coverage across all ages and risk groups [2]. Fourth, in our model vaccination is taken at random, while in practice strong individual-specific preferences are present [3, 2]. Together, these factors could make a large vaccination-driven postponement of infections from midlife into very old age less likely. At the same time, two points argue for taking the mechanism seriously. The first is that the development of novel influenza vaccines is aimed at achieving higher effectiveness, broader strain coverage, and longer duration of protection [11, 18, 35]. If in practice such novel vaccines happen to provide strong but short-lived protection, the conditions for infection age-shifting become more relevant. The second is that, for pathogens with stable antigenic targets and long-lasting natural immunity, repeated vaccination could plausibly create a larger difference between vaccine and infection immunity durations. This, in turn, could bring the harmful regime closer. Our results therefore function as a cautionary benchmark. As vaccines continue to evolve, vaccination programs must consider not only the direct protection of vaccinated individuals, but also the life-course redistribution of infection risks.

Our main analyses assumed an infection fatality pattern that increases monotonically with age, which is appropriate for SARS-CoV-2 and influenza A when focusing on fatal outcomes [8, 2, 27]. However, it does not capture pathogens with pronounced U-shaped severity profiles, where the overall disease burden is focused in infants and older adults (notably pertussis and RSV). Introducing a U-shaped severity function could strengthen the case for protecting against early-life infections, either directly via infant vaccination or indirectly via maternal immunization and cocooning. As a result, it could also alter the trade-off between early and late vaccination, favoring early vaccination to prevent severe pediatric disease. Extending the current model to incorporating such severity profiles is therefore a natural next step for pathogens such as RSV and pertussis, where new vaccination and monoclonal antibody strategies are now being deployed in pregnant women and infants.

Several limitations should be kept in mind. We used a stylized demographic model with fixed births and no migration, assumed an all-or-nothing vaccine that fully prevents infection, and did not include partial protection against infection, reductions in severity conditional on infection, age-dependent vaccine take (see for a review, [15]), or vaccine effectiveness depending on vaccination history [31, 17]. All of these factors are relevant for influenza A, and most are also relevant for RSV and SARS-CoV-2. Our analyses further assume infection fatality risks that increase steeply with age, which is appropriate for pathogens such as influenza A and SARS-CoV-2 but not for directly for infections where severe outcomes are concentrated in infancy or childhood. We also focused on long-run averages rather than short-run program transitions. These simplifications allowed us to focus on the mechanisms by which vaccination changes the age distribution of infections and mortality. Nonetheless, they should be addressed in future work aimed at pathogen-specific policy evaluation.

An important implication of our results is that adult vaccination should be assessed in a multi-decade perspective, across the remaining life course of all individuals in the population. Vaccine effectiveness analyses increasingly recognize that protection (against symptomatic outcomes) depends on prior infection and vaccination history, immune imprinting, and age-related immune declines (reviewed in [31, 17]). However, studies into the long-term population-level impacts are still lacking. Our analyses have provided first steps in this direction. Our main conclusion is not that vaccination is harmful, but that vaccination strategies for older adults must be designed with explicit attention to life-history effects. As adult immunization programs expand and vaccine formulations continue to evolve such life-history considerations will become increasingly important to for efforts to minimize the disease burden in older adults.

## Methods

### Model equations

We consider a time- and age-dependent SIRV transmission model with non-permanent immunity. Individuals are classified as susceptible (*S*), infected and infectious (*I*), immune by natural infection (*R*), or immune by vaccination (*V*). To enable realistic distributions of immune durations we include multiple *R* and *V* compartments, *n*_*R*_ and *n*_*V*_, such that the overall immune durations are Erlang distributed [6]. The model is given as a set of partial differential equations for the densities *S*(*t, a*), *I*(*t, a*), *R*_*i*_(*t, a*) and *V*_*j*_(*t, a*), depending on time *t* and age *a*. Let 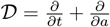 denote the age–time derivative operator. Then,

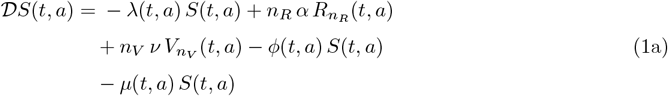

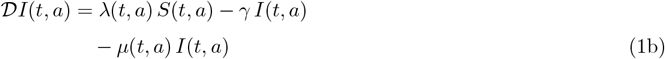

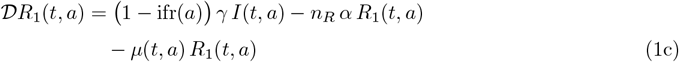

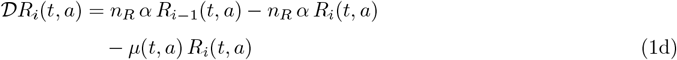

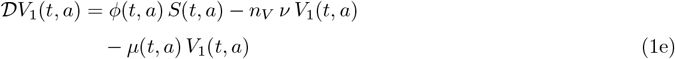

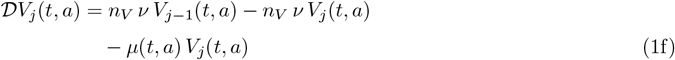

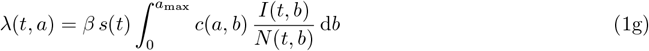

with *i* = 2, …, *n*_*R*_ and *j* = 2, …, *n*_*V*_. Notice that the mean durations of natural and vaccine-induced immunity are given by 1*/α* and 1*/ν*, respectively. Further,

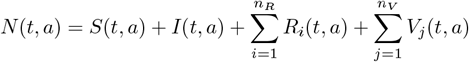

denotes the population density at age *a* and time *t*. The force of infection *λ*(*t, a*) in the above formulation assumes frequency-dependent transmission. For convenience, we scale the contact function *c*(*a, b*) such that, after discretization, its dominant eigenvalue equals 1. Hence, if we take the seasonal forcing multiplier *s*(*t*) ≡ 1, the basic reproduction number is given by ℛ_0_ = *β/γ*.

In the above formulation, vaccination moves individuals from *S* to *V*_1_ at a rate *ϕ*(*t, a*). Individuals in *R* and *V* are assumed to be fully protected against infection until immunity wanes. Hence, this implies all-or-nothing protection whereby individuals are either fully immune or fully susceptible. Infection-induced mortality is implemented by assuming that a fraction ifr(*a*) of individuals leaving the infectious compartment die (and a fraction (1 − ifr(*a*)) enters the *R*_1_ compartment upon recovery).

All newborn individuals enter the population in the susceptible compartment, i.e. *S*(*t*, 0) = *N*_0_ and all other boundary conditions at *a* = 0 are zero. Simulations are initialized at a demographic equilibrium implied by the 2025 mortality rates (*µ*(2025, *a*)) and run through a 150-year burn-in period with a small initial infection seed to reach a stable compartmental age distribution. Table 1 provides an overview of the main model parameters and functions, with default values.

**Table 1.** Model parameters.

| Parameter | Explanation | Default value |
| --- | --- | --- |
| $N_0$ | birth cohort size | 200,000 yr<sup>-1</sup> |
| $\beta$ | transmission rate parameter | 150 yr<sup>-1</sup> |
| $1/\gamma$ | infectious period | 0.02 yr |
| $\mathcal{R}_0$ | basic reproduction number | $\mathcal{R}_0 = \beta/\gamma = 3$ |
| $s(t)$ | seasonal forcing multiplier | $1 + f \cos(2\pi t)$ |
| $f$ | amplitude of seasonal forcing | 0.05 |
| $c(a, b)$ | person ($a$) to group ($b$) contact rates | [39] |
| $\lambda(t, a)$ | force of infection | Eq. 1 |
| $1/\alpha$ | mean duration of natural immunity | $\mathcal{U}(2, 7)$ yr (influenza A) |
| $1/\nu$ | mean duration of vaccine immunity | 0.5 – 5 yr (influenza A) |
| $n_R$ | number of immune compartments | 2 |
| $n_V$ | number of vaccinated compartments | 2 |
| $a_{\max}$ | maximum age in the model | 120 yr |
| $\phi(t, a)$ | vaccination rate | see text |
| $\mu(t, a)$ | mortality rate | see text |
| $\text{ifr}(a)$ | infection fatality probability | see text |

### Demography

In Eq. 1, the natural mortality rate (i.e. non–infection-related) is given by the log-linear function

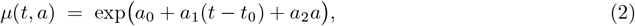

where *t* denotes time in years, *t*_0_ is the reference year, *a* is age in years. We take *t*_0_ = 2020 as the reference year and assume a constant birth cohort size, *N*^∗^(*t*, 0) = *N*_0_. Let *N*^∗^(*t, a*) denote the population density by age in the absence of infection-induced mortality. Consistent with the age–time derivative operator *D* in (1), aging of a cohort with natural mortality only can be described by

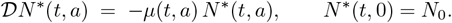

Solving this yields

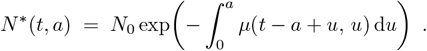

For the log-linear mortality rate (2), this can be evaluated explicitly, giving

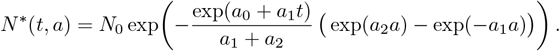

In the full transmission model, the total population density is given by 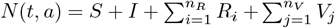, and infection-induced mortality removes individuals from the population at a rate ifr(*a*)*γI*(*t, a*). For the demographic calculations above, we neglect this term because it does not materially affect the population age composition in the scenarios considered.

In our analyses, the parameters (*a*_0_, *a*_1_, *a*_2_) were obtained by fitting Eq. (2) to age- and time-specific mortality rates for the Netherlands (see Table 1 and data sources described in the main text), yielding *a*_0_ = −5.054, *a*_1_ = −0.006121, and *a*_2_ = 0.04695. In the following we use (2) to calculate (remaining) life expectancy and (expected) years-of-life-lost (YLL).

### Contact patterns

Contact patterns are estimated from a human contact study carried out in the Netherlands in 2006-2007 [24]. In this study a total 825 participants recorded characteristics of 11,225 contacts with unique individuals, and a hierarchical Bayesian model was fitted to the data to estimate smoothed person-to-person contact rates [39]. In these analyses the maximum age was restricted to 80 years as only few contacts were with persons older than 80 years, and even fewer were recorded with persons older than 100 years. Here, we have rerun the analyses using a maximum age of 120 years. Thus, estimates of contact rates in older persons are based on a small number of persons and for a considerable part extrapolated from information contained in data from younger age groups. Figure 7 shows the estimated symmetrical person-to-person contact rate matrix **C**_sym_. Our transmission model uses a frequency-dependent transmission term, and the contact matrix in our model is based on person to group contact rate matrix **C** = **C**_sym_diag(**w**), where **w** is the age composition of the population at time *t* = 0. Notice that this implies that the age-mixing pattern is calibrated to the baseline demographic composition (at *t* = 0) and is held fixed thereafter. In particular, we do not update **w** over time, so demographic change does not induce a time-varying contact matrix [39, 1]. This is common practice in infectious disease modeling.

**Figure 7.**
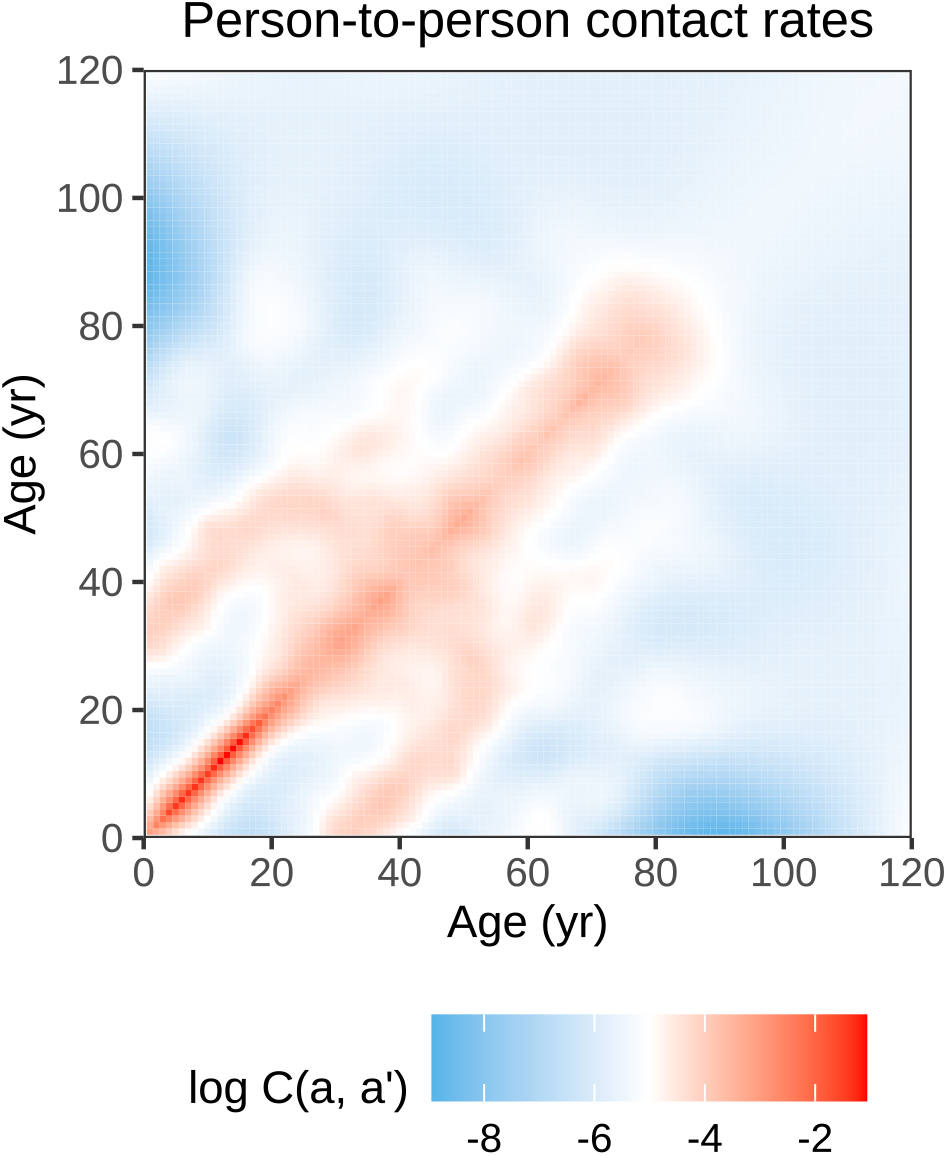
Person-to-person contact matrix as estimated from a contact study in the Netherlands in 2006-2007. The matrix has been normalized so that its dominant eigenvalue equals 1. Notice the strong age-assortative patterns, indicating that persons with similar ages have high number of contacts. Also note the sub- and super-diagonal bands indicating high contact rates between parents and children. See for details [39].

### Seasonal forcing

Non-forced SIR-type transmission models with demographic turnover have the inherent tendency of approaching an endemic equilibrium, where the rates of infection, recovery, and replenishment of susceptible individuals balance out, leading to a stable equilibrium [9]. For respiratory pathogens this is unrealistic as it is known that transmissibility is inherently higher in winter than in summer. Also, this prevents the recurrent epidemics that are typical for respiratory infections. Here we incorporate simple sinusoidal seasonal forcing strength of 5% [36, 25]. Specifically, if we denote by *f* the forcing strength, the forcing function is given by

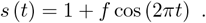

### Infection-induced mortality and years-of-life-lost (YLL)

Infection-induced mortality is obtained by linking the modeled incidence of infection (or, more precisely, incidence of exit from infection) at time *t* and age *a* to the age-specific infection fatality probability ifr(*a*) (Table 1). We use an exponentially increasing function of age,

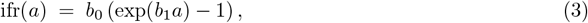

with *b*_0_ = 1.86 *×* 10^−5^ and *b*_1_ = 0.07. This choice yields ifr(0) = 0, ifr(80) ≈ 0.005, ifr(90) ≈ 0.01 and ifr(100) ≈ 0.02. These values are broadly in line with age patterns reported for seasonal influenza mortality and severity in high-income settings [38, 33, 3].

With the infection fatality probability at hand, the age-specific infection-induced mortality incidence is given by

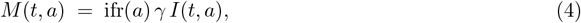

where ifr(*a*) converts infections into deaths. Age-specific years of life lost are given by

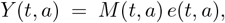

with *e*(*t, a*) the remaining (cohort) life expectancy at time *t* and age *a*.

Total mortality and YLL at time *t* are obtained by integrating over age,

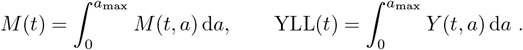

In the influenza A scenarios we fix the (background) mortality schedule to 2025 and 2050 by evaluating it at *µ*(2025, *a*) or *µ*(2050, *a*), while deriving cohort life expectancy *e*(*t, a*) from that point onward.

### Vaccination and duration of immunity

Throughout we assume vaccination scenarios in which a fixed fraction of the population is vaccinated and fully protected against infection. Hence, we assume an all-or-nothing vaccine. Three scenarios are considered. To set the stage, we first consider a non-forced transmission model (i.e. *s* (*t*) ≡ 1) and a vaccine that provides protection for half the duration of natural infection (5, 10, 20, and 50 years, Figure 3).

In this scenario we introduce the vaccination persons reach a given age, and the vaccination rate *ϕ*(*t, a*) becomes independent of time. Here, the focus is on the vaccination strategy that minimizes mortality or YLL in 2025 or 2050. The second scenario extends the first but with repeated vaccination and seasonal forcing (mimicking influenza A). In this scenario vaccination is given to all persons from a certain age onward in October and November. In both scenarios, the vaccination rate is tuned such that the required effective coverage *c* is reached within a campaign window of duration Δ_vac_ years, i.e. 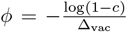. Immunity durations after infection and vaccination are represented by Erlang-distributed durations (with *n*_*R*_ = *n*_*V*_ = 2). Hence, standard deviations of these distributions are 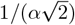 for infection-induced immunity and 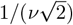 for vaccine-induced immunity. While vaccine induced is assumed to be fixed (0.5 − 5 years), the duration of natural immunity changes from season to season by (unpredictable) antigenic drift of the virus, and is uniformly distributed between 2 and 7 years (Table 1).

### Numerical integration

The model was implemented in Julia (version 1.11.4). We solved the time–age structured PDEs using the method of lines, discretizing age (*a* ∈ [0, 120)) on a uniform grid with Δ*a* = 0.1 years (yielding *n*_*a*_ = 1200 bins), and the age terms were approximated by a first-order upwind scheme. Inflow by births into the population at *a* = 0 was implemented through the susceptible class. This yields a set of coupled ODEs of size 6*n*_*a*_. We integrated the ODEs with `DifferentialEquations.jl` using the adaptive Runge–Kutta solver `Tsit5()` (tolerances `reltol=1e-4`, `abstol=1e-6`), while enforcing non-negativity caused by numerical integration errors with `DiffEqCallbacks.jl`. Together, this ensured precise approximations to the PDE model with reasonable step-sizes. For parameter sweeps we saved the solution on regular parameter grids and parallelized runs using `ThreadsX.jl`. Postprocessing of all output was done in R (version 4.5.1). All code and data required to reproduce the figures is available at github.com/mvboven/ageing.

## Data Availability

https://github.com/mvboven/ageing

## Acknowledgements

This work was supported by the Innovative Medicines Initiative 2 Joint Undertaking under Grant Agreement 101034339 and ZonMw project 16230396.

## Author contributions

Conceptualization: all authors. Analyses: MvB. Writing: all authors.

## Data and code availability

All code and data required to reproduce the figures are available at https://github.com/mvboven/ageing.

